# Acute kidney injury in patients with severe COVID-19

**DOI:** 10.1101/2020.08.28.20167379

**Authors:** Gustavo A. Casas-Aparicio, Isabel León-Rodríguez, Mauricio González-Navarro, Claudia Alvarado-de la Barrera, Santiago Ávila-Ríos, Amy Peralta-Prado, Yara Luna-Villalobos, Alejandro Velasco-Morales, Natalia Calderón-Dávila

**Affiliations:** Centro de Investigación en Enfermedades Infecciosas, Instituto Nacional de Enfermedades Respiratorias Ismael Cosío Villegas; Departamento de Epidemiología, Instituto Nacional de Enfermedades Respiratorias Ismael Cosío Villegas; Resident doctor at the Instituto Nacional de Enfermedades Respiratorias Ismael Cosío Villegas

**Keywords:** Acute kidney injury, AKI, acute renal failure, COVID-19, SARS-CoV-2.

## Abstract

**Introduction:** Some patients with COVID-19 pneumonia present systemic disease involving multiple systems. There is limited information about the clinical characteristics and events leading to acute kidney injury (AKI). We described the factors associated with the development of AKI and explored the relation of AKI and mortality in Mexican population with severe COVID-19.

**Methods:** We retrospectively reviewed the medical records of individuals with severe pneumonia caused by SARS-CoV-2 hospitalized at the largest third-level reference institution for COVID-19 care in Mexico between March and April 2020. Demographic information, comorbidities, clinical and laboratory data, dates of mechanical ventilation and hospitalization, mechanical-ventilator settings and use of vasoactive drugs were recorded.

**Results:** Of 99 patients studied, 58 developed AKI (58.6%). The group with AKI had higher body mass index (p=0.0003) and frequency of obesity (p=0.001); a higher requirement of invasive mechanical ventilation (p=0.008) and vasoactive drugs (p=0.004); greater levels of serum creatinine (p<0.001) and D-dimer on admission (p<0.001); and lower lymphocyte counts (p=0.001) than the non-AKI group. The multivariate analysis indicated that risk factors for AKI were obesity (adjusted hazard ratio (HR)=2.71, 95% confidence interval (CI)=1.33-5.51, p=0.005); higher serum creatinine (HR=1.44, CI=1.02- 2.02, p=0.035) and D-dimer levels on admission (HR=1.14, CI=1.06-1.23, p<0.001). Inhospital mortality was higher in the AKI group than in the non-AKI group (65.5% vs. 14.6%; p=0.001).

**Conclusions:** AKI was common in our cohort of patients with severe COVID-19 and it was associated with mortality. The risk factors for AKI were obesity, elevated creatinine levels and higher D-dimer levels on admission.

## INTRODUCTION

In December 2019, a series of pneumonia cases of unknown cause emerged in Wuhan, Hubei Province, China, with clinical presentations resembling viral pneumonia [1]. The pneumonia spread quickly to other provinces of China and overseas. A novel coronavirus was identified by the Chinese Center for Disease Control and Prevention (CDC) from the throat swab sample of a patient and was provisionally named 2019-nCoV by the World Health Organization (WHO) [2]. Based on phylogeny, taxonomy and established practice, the International Committee on Taxonomy of Viruses renamed the virus as Severe acute respiratory syndrome coronavirus-2 (SARS-CoV-2) [3]. WHO subsequently declared coronavirus disease 2019 (COVID-19) a public health emergency of international concern [4]. COVID-19 is primarily manifested as a respiratory tract infection, but emerging data indicate that it should be regarded as a systemic disease involving multiple systems, including cardiovascular, respiratory, gastrointestinal, neurological, hematopoietic, immune and renal [5, 6, 7, 8]. Of note, after lung infection, the virus may enter the blood, accumulate in the kidney and cause damage to resident renal cells, with a significantly higher risk for in-hospital death [8]. Thus, understanding how the kidney is affected by SARS-CoV-2 is particularly relevant. The incidence of acute kidney injury (AKI) in hospitalized patients with COVID-19 varies across populations, but a large multicenter retrospective cohort study in New York reported AKI in 37% of hospitalized patients, and 35% of those died [9]. AKI initiation coincides with the development of Acute Respiratory Distress Syndrome (ARDS), and these alterations are typical of patients progressing to the most severe stage of illness involving extra-pulmonary systemic hyperinflammation [10]. The National Institute of Respiratory Diseases (INER) is the largest third-level national referral center for COVID-19 in Mexico City. Since early January 2020, this institution was gradually repurposed for the treatment of patients with COVID-19 exclusively. Since February 28, 2020, when the first Mexican patient was diagnosed with COVID-19, a high proportion of critically ill patients have been admitted at the INER. The aim of this study was to describe the factors associated with the development of AKI and explore the relation of AKI and mortality in the Mexican population with severe COVID-19.

## METHODS

### Study population

The study was conducted at the National Institute of Respiratory Diseases (INER), the largest third-level institution designated by the Mexican Government for COVID-19 care. All medical records of individuals with severe pneumonia caused by SARS-CoV-2 hospitalized at the INER between March and April 2020 were retrospectively reviewed. The Institutional Review Board approved the study and waived the requirement for informed consent due to the retrospective design of the stud (Approval No. C39-20). We included individuals with diagnosis of severe pneumonia caused by SARS-CoV-2, confirmed by real-time reverse transcription-polymerase chain reaction (rRT-PCR); 18 years of age or older; with no history of chronic kidney disease (CKD); and ratio of partial arterial oxygen pressure/inspired oxygen fraction (PaO_2_/FiO_2_) <300 mm Hg on admission. Pregnant women were not included in the study. Patients with incomplete clinical records were excluded. Patients with SARS-CoV-2 severe pneumonia were defined as those with clinical data of respiratory distress, bilateral alveolar opacities in 2 or more lobes, a ratio of PaO_2_/FiO_2_ < 300 mm Hg and a positive result for SARS-CoV-2-rRT-PCR assay [11] in nasopharyngeal swab. Prone ventilation was used for the treatment of ARDS as a strategy to improve oxygenation when traditional modes of ventilation failed.

The primary outcome was the development of AKI. The secondary outcome was 30-day mortality in the group with AKI and the group without AKI. Recorded variables included demographic and anthropometric variables, symptoms, comorbidities, treatments, critical care variables, blood chemistry, blood count, initiation and termination dates of mechanical ventilation, days in hospital, initial mechanical-ventilator settings, use of vasoactive drugs and outcomes.

### Acute kidney injury

Diagnosis of AKI was based on a rapid reduction of kidney function, defined by an absolute increase in adjusted serum creatinine (sCr)> 0.3 mg/dL within 48 hours compared to baseline; or a sCr increase of 1.5-fold compared to baseline within 7 days. AKI stage was defined according to the Kidney Disease Improving Global Outcomes (KDIGO) staging system [12]. AKI stage 1 corresponded to a sCr increase of 1.5±1.9 times baseline; AKI stage 2 corresponded to a sCr increase 2.0±2.9 times baseline; and AKI stage 3 corresponded to a sCr increase of ≥3 times baseline or initiation of renal replacement therapy (RRT). Patients were stratified according to the highest AKI stage attained during hospital stay. As none of the patients had pre-hospital baseline sCr measurements available, the baseline value was retrospectively adjusted to the median sCr value from the entirety of the hospitalization period (this value was chosen, rather than the nadir or final creatinine to provide a more conservative estimate to adjudicate AKI) [8]. The median serum creatinine value during hospitalization was used for diagnosis of AKI. The urine output criterion was not used for diagnosis of AKI since nursing records were out of reach, in COVID-19 areas.

### Furosemide Stress Test

Patients with AKI underwent the furosemide stress test for prediction of AKI outcome. The test was performed by administrating 1 mg/kg of furosemide i.v. or 1.5 mg/kg if the patient received furosemide within the preceding 7 days, followed by observation of the urinary output in the first 2 hours. The result was considered positive if the patient urinated more than 200 ml per hour, in the following 2 hours after furosemide administration.

### Statistical Analysis

We performed descriptive statistics including means and standard deviations for normally distributed continuous variables, medians and interquartile ranges for non-parametric distributions, and proportions for categorical variables. Comparisons of the AKI vs. the non-AKI groups were made using Fisher’s exact test for categorical variables and U Mann-Whitey for continuous variables. Comparisons across AKI stages were made using Kruskal-Wallis rank sum test. All statistical tests were two-sided and p values <0.05 were considered statistically significant. We performed sensitivity analysis for missing data. Cox proportional Hazard regression analysis was used to identify association between relevant covariates and AKI. No violations of the assumptions were detected. The multivariate model was built using a stepwise procedure, and variables were entered into the models when they reached a cutoff p<0.02 in the univariate analysis. We also performed the Kaplan-Meier survival analyses for the time to death, comparing the group with AKI vs. the non-AKI group. Statistical analyses were performed using R version 3.6.3.

## RESULTS

### Characteristics of study population

During the period between March 1^st^, 2020 and April 30, 2020, a total of 280 individuals were admitted at the INER due to suspected COVID-19. Of those, 12 died during the first 48 hours and 78 had a negative result for the SARS-CoV-2 rRT-PCR test. Therefore, we reviewed the clinical files of 190 individuals. Of those, 12 had pneumonia due to other causes; 60 had incomplete clinical files; and 19 were transferred to other hospitals. We thus included 99 individuals in the study (Figure 1). Of those, 74 were male (74.7%); the median age was 52.9 years (SD±13.27); 30 had hypertension (29.7%); 27 had diabetes (26.7%); and 56 had obesity (55.4%; Table 1). On admission, 82.8% had a positive result for SARS-CoV-2-rRT-PCR, but all patients had a positive result when the test was performed for the third time. Sixty-five patients (64.3%) required invasive mechanical ventilation (IMV). The median positive end-expiratory pressure (PEEP) level was 11.5 cm H_2_O (SD +2.4); and 28 required prone ventilation due to refractory hypoxemia. Forty-nine patients (48.5%) required vasoactive drugs on admission; and overall mortality was 44.4%. All patients had ground glass opacities; 85.2% had crazy paving pattern; 94.3% had consolidation; 5.6% had pleural effusion; 79.5% had bronchiectasis; 54.5% had atelectasis; 59% had peripheral distribution; and 39.7% had central and peripheral distribution.

**Table 1.**
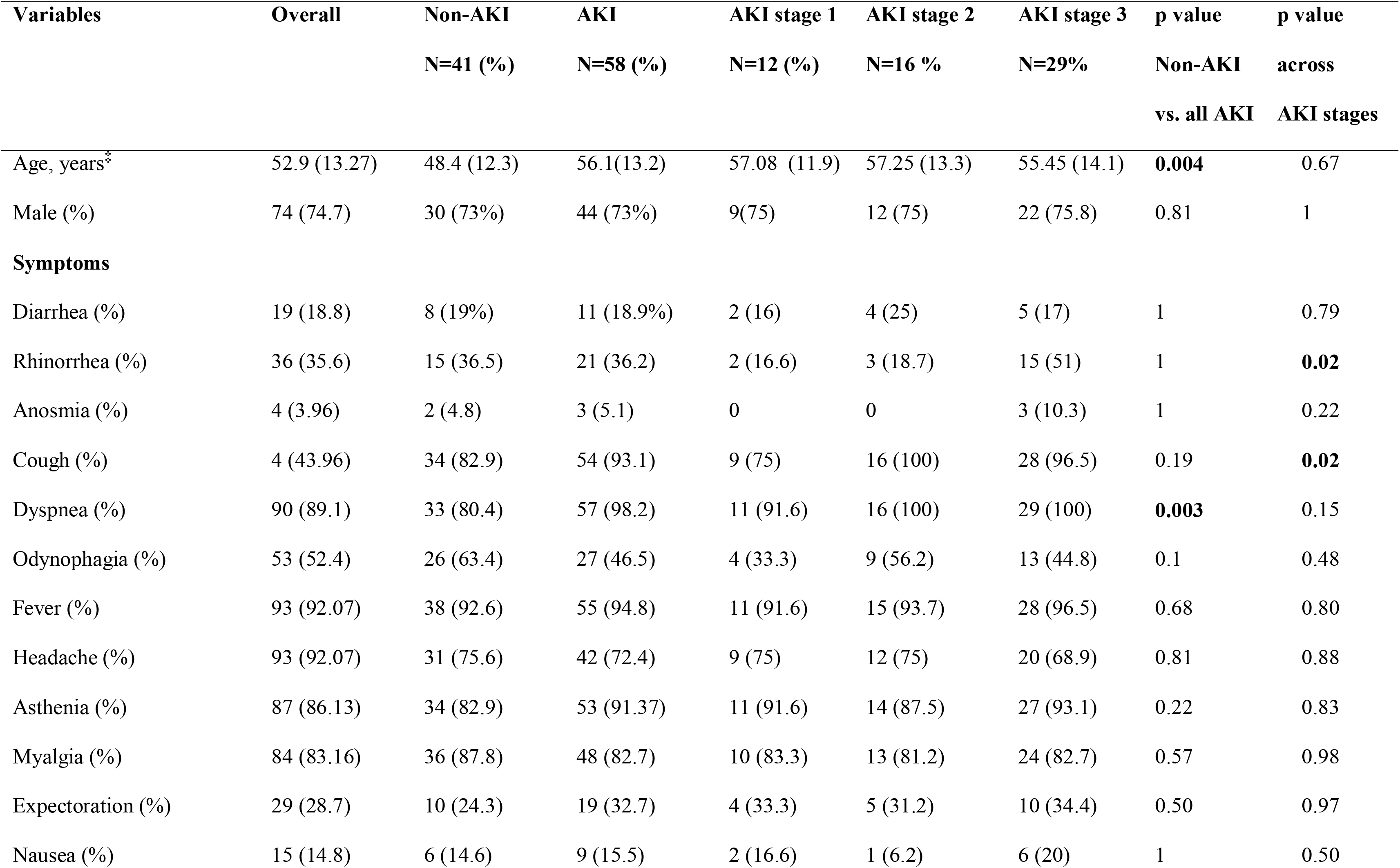
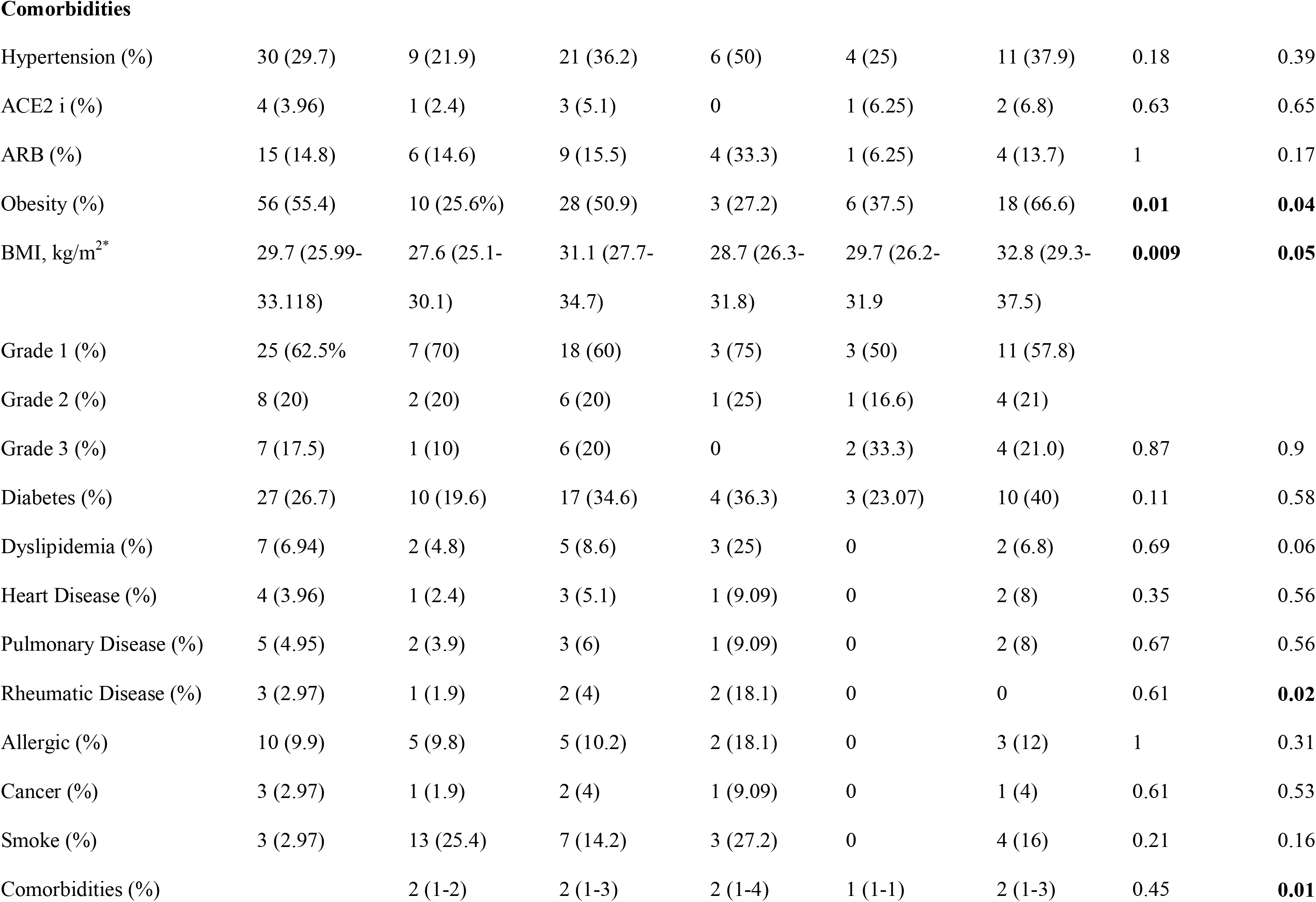
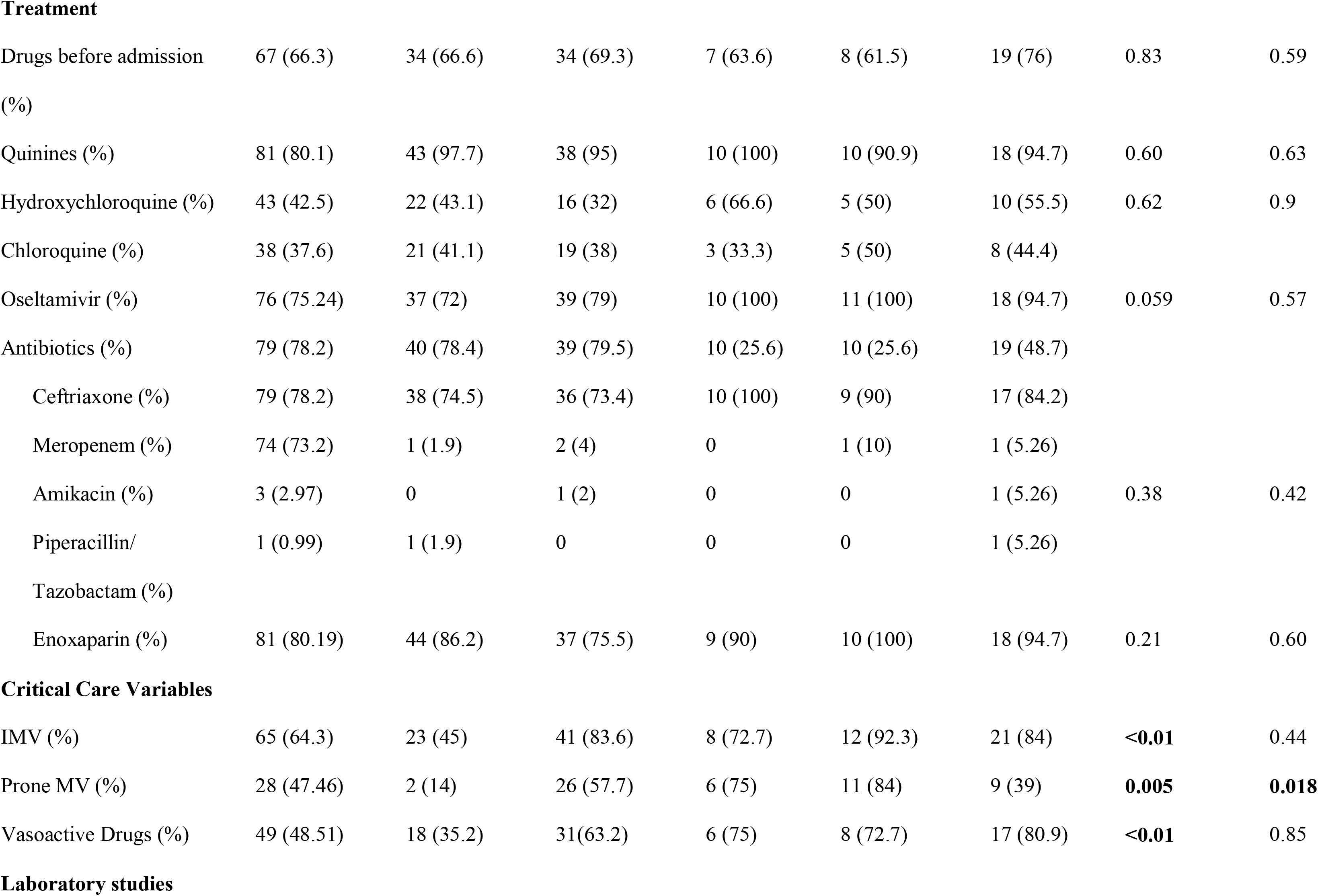
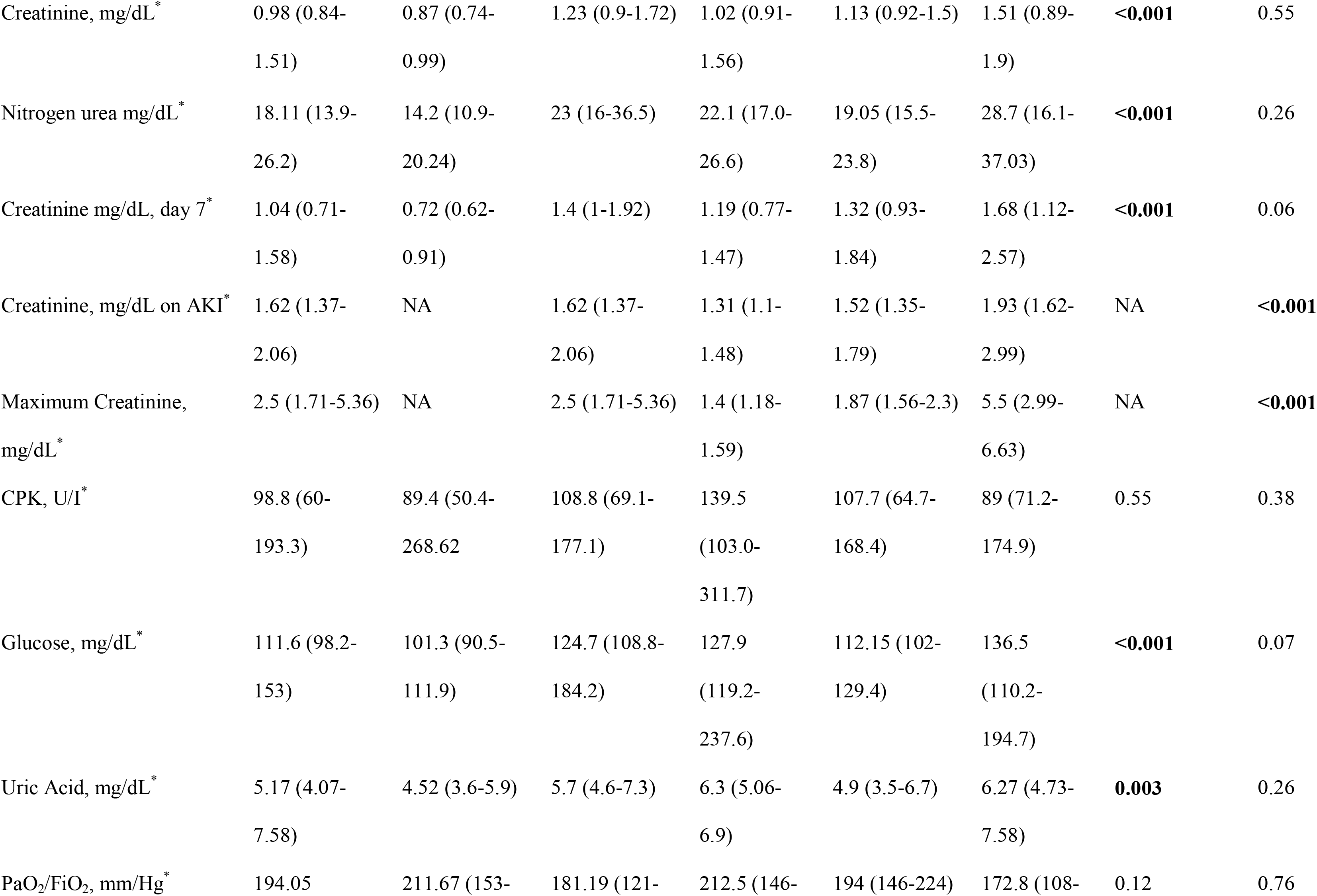
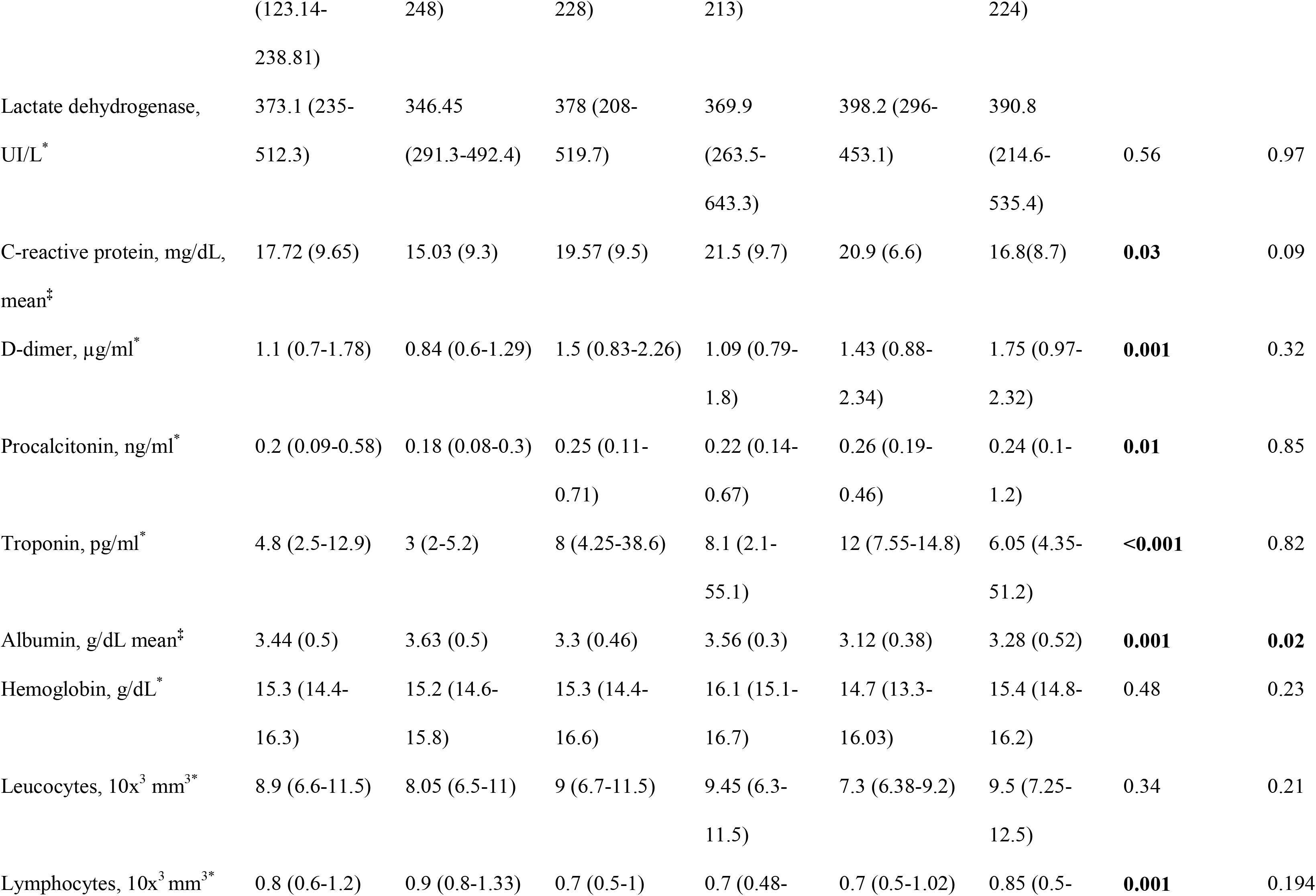
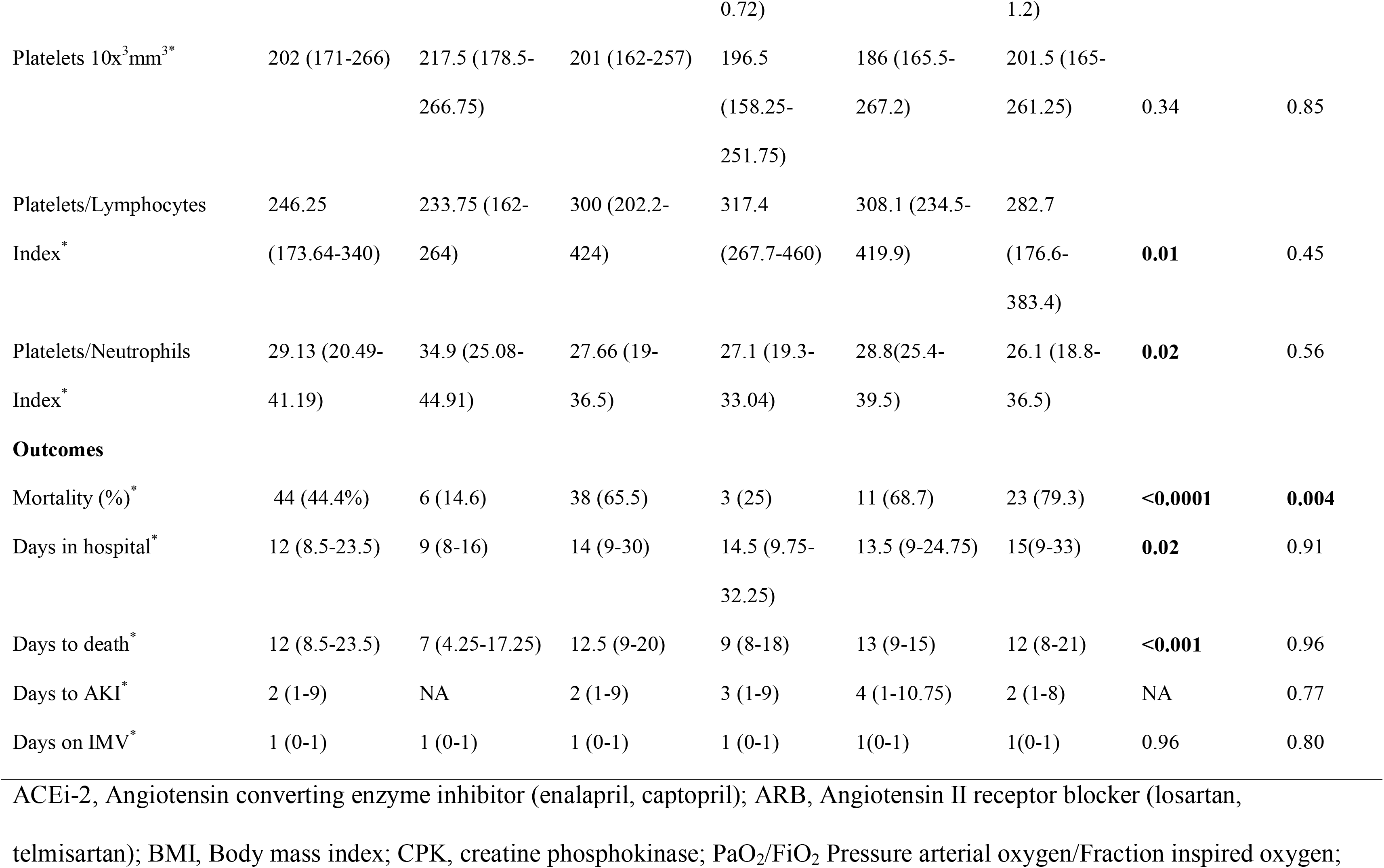
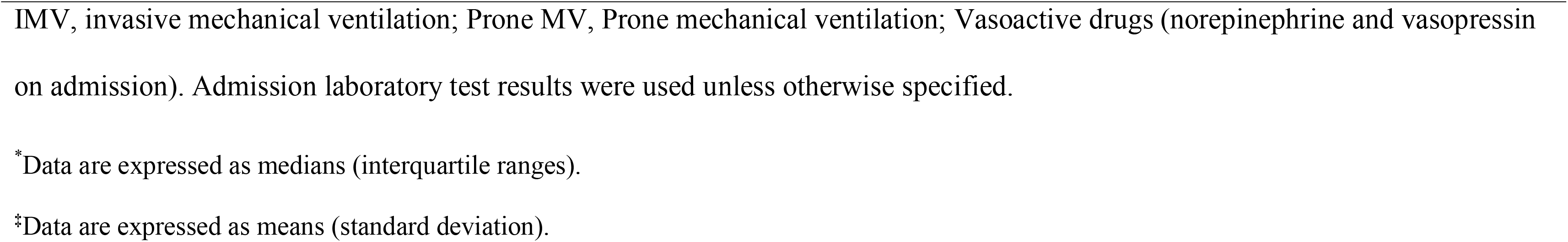
Characteristics of the study population.

**Figure 1.**
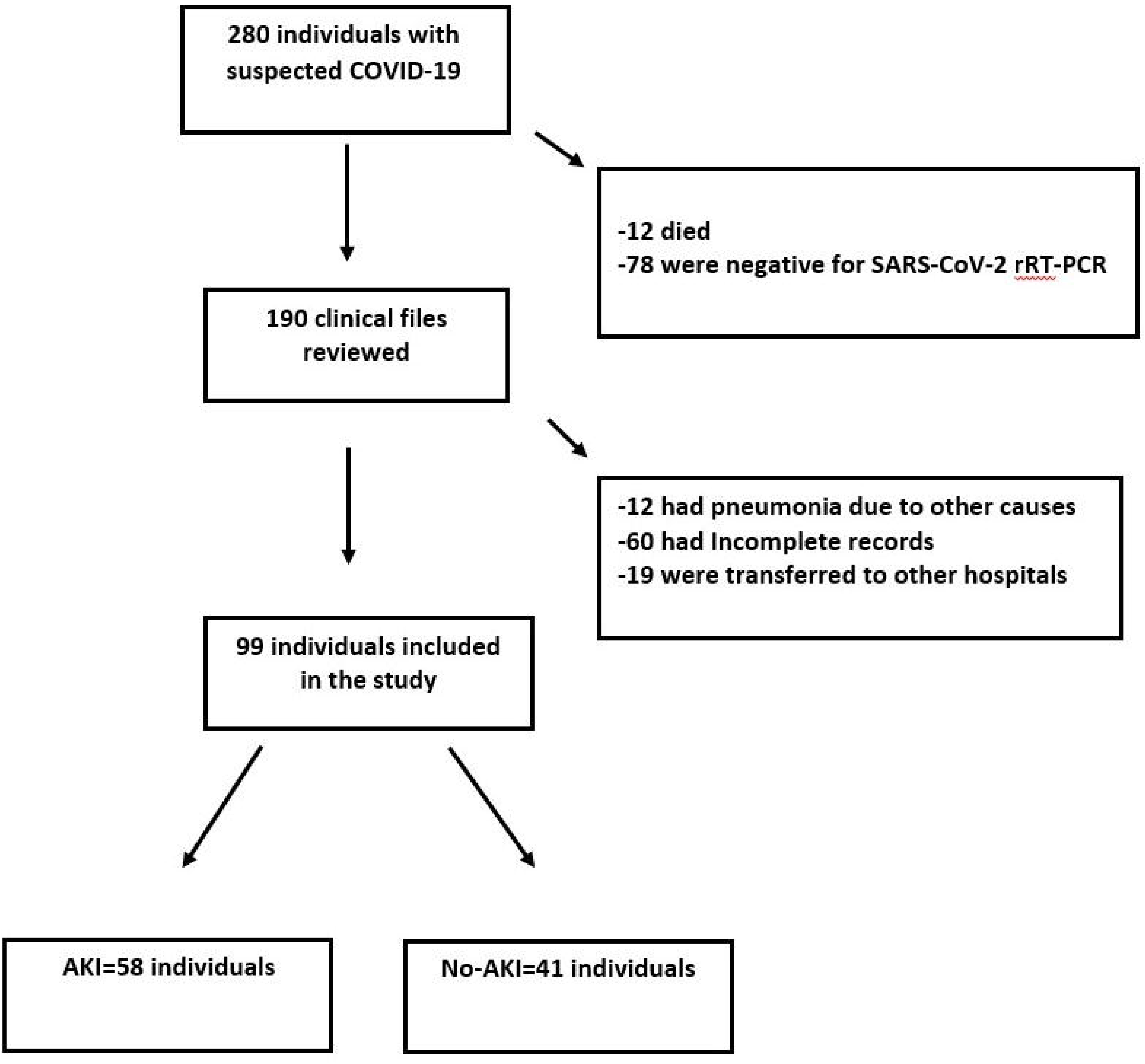
Study diagram. Numbers of individuals assessed for eligibility and individuals included in the study.

### Acute Kidney Injury

Fifty-eight patients developed AKI (the AKI group) and 41 individuals did not develop AKI (the non-AKI group, Table 1). Of those, 12 had AKI stage 1 (21.1%); 16 had AKI stage 2 (28.1%); and 29 had AKI stage 3 (50.9%). Forty-one patients of the AKI group (83.6%) required IMV, while 23 patients of the non-AKI group (45%) required IMV (p=0.01). Twenty-six patients of the AKI group required prone mechanical ventilation (57.7%), compared with 2 patients (14%) in the non-AKI group (p=0.005). On admission, 31 patients of the AKI group required vasoactive drugs (63.2%), compared with 18 patients (35.2%) of the non-AKI group (p=0.001). In-hospital mortality was significantly higher in the AKI group (38 patients in the AKI group (65.5%) vs. 6 patients (14.6%) in the non-AKI-group; (p=0.001; Figure 2). In-hospital mortality was significantly higher in patients with AKI 3 (79.3%) and AKI 2 (68.7%) compared with those with AKI 1 (25%); p=0.004). The body mass index (BMI) was significantly higher in the AKI group (31.1 kg/m^2^ vs. 27.6 kg/m^2^ in the non-AKI group; p=0.009). Obesity was more frequent in the AKI group (28 patients, 50.9%) than in the non-AKI group (10 patients, 25.6%; p=0.01). The median time to AKI development was 6.5 days (SD+8.33). Thirty-nine percent of the patients with AKI had a positive result for the furosemide stress test. A total of 11 patients (22.4%) required renal replacement therapy (RRT). Of those, 5 used continuous RRT, 3 used intermittent hemodialysis (IHD), and 3 used prolonged intermittent renal therapies (PIRT). The median time to RRT initiation after AKI initiation was 4.18 days (SD±3.8) and median time under RRT was 4.29 days (SD+ 2.82). Five patients died after 30 days of follow-up, 2 of them had been discharged due to clinical improvement and 3 were hospitalized.

**Figure 2.**
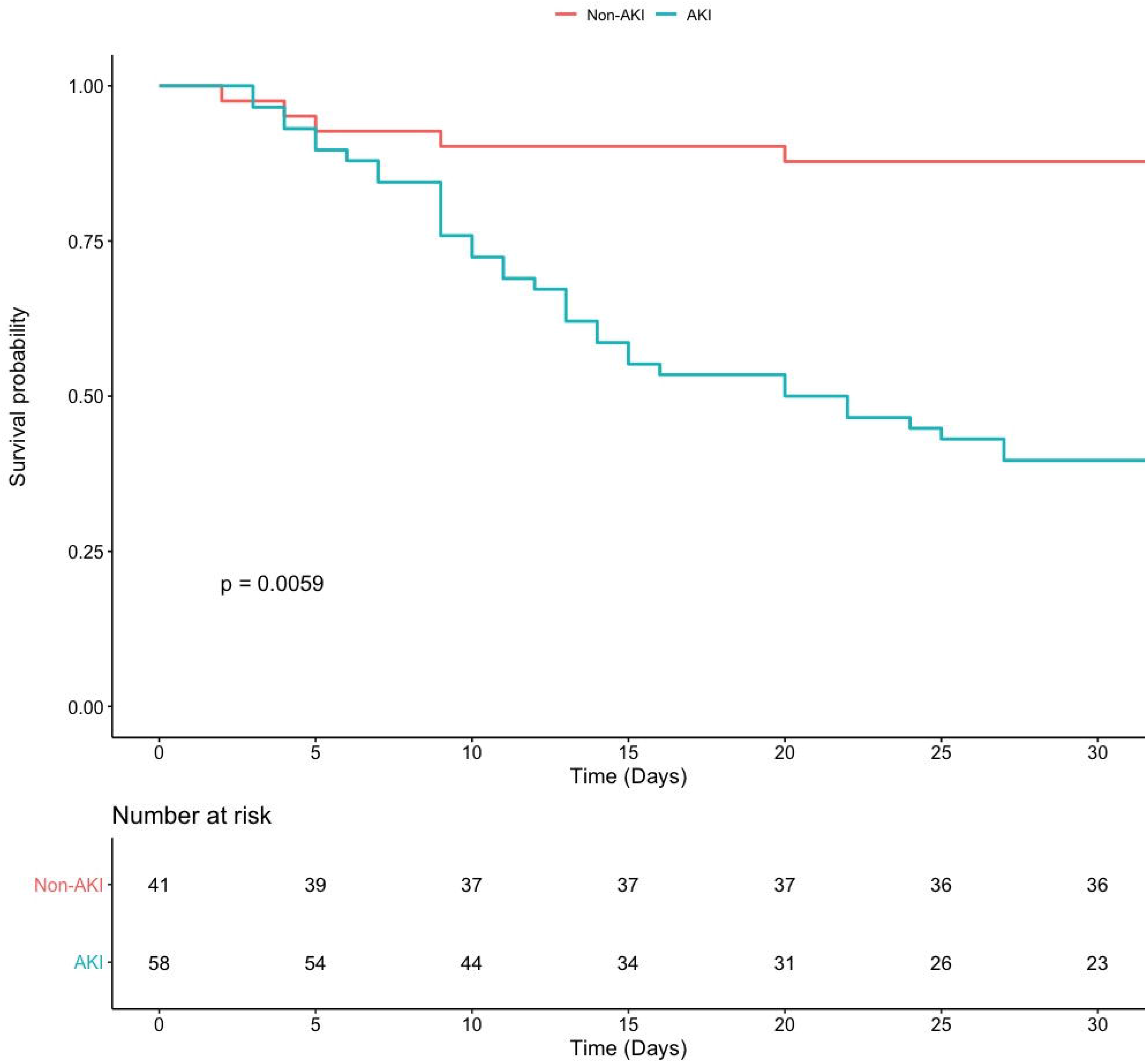
Kaplan-Meier survival curves. Time to death for the AKI group (blue line), and the non-AKI group (red line) during a follow-up period of 30 days. Time 0 corresponded to hospital admission.

### Inflammation markers and AKI

Some inflammation markers were higher in the AKI group, including D-dimer: 1.5 μg/ml (interquartile range (IQR), 0.83-2.26) in the AKI group vs. 0.84 μg/ml (IQR, 0.6-1.29) in the non-AKI group, p=0.001; procalcitonin: 0.25 ng/ml (IQR, 0.11-0.71) in the AKI group vs 0.18 ng/ml (IQR, 0.08-0.3) in the non-AKI group, p=0.01; and troponin: 8 pg/ml (IQR, 4.25-38.6) in the AKI group vs. 3 pg/ml (IQR, 2-5.2) in the non-AKI group, p<0.001. Lymphocytes were decreased in the AKI group: 0.7 10x^3^ mm^3^ (IQR, 0.5-1) compared to the non-AKI group: 0.9 10x^3^ mm^3^ (IQR, 0.8-1.33, p=0.001; Table 1).

### Risk factors for acute kidney injury

The univariate analysis indicated that patients with AKI had a higher BMI (unadjusted hazard ratio (HR)=1.1, 95% confidence interval (CI) 1-1.1, p=0.0003); higher frequencies of obesity (BMI>30 kg/m^2^, HR=2.4, 95% CI 1.4-4.1, p= 0.001); a higher requirement of IMV (HR=0.4, 95% CI=0.2-0.79, p=0.008); a higher use of vasoactive drugs (HR=0.46, 95% CI=0.27-0.79, p=0.004); higher creatinine levels on admission (HR=1.7, 95% CI=1.4- 2.0, p<0.001); lower lymphocyte counts (HR=0.39, 95% CI=0.19-0.78, p=0.008); higher D-dimer levels on admission (HR=1.1, 95% CI=1.1-1.2, p<0.001); and lower albumin levels HR=0.48, 95% CI=0.28-0.88, p=0.007) than non-AKI patients.

After adjusting for possible confounding variables, the multivariate analysis indicated that risk factors for AKI were obesity (adjusted HR=2.71, 95% CI=1.33-5.51, p=0.005); higher serum creatinine (HR=1.44, CI=1.02-2.02, p=0.035) and D-dimer levels on admission (HR=1.14, CI=1.06-1.23, p<0.001, Table 2).

**Table 2.** Risk factors for Acute Kidney Injury.

| **Variables** | **Unadjusted (HR 95% CI)** | **p value** | **Adjusted (HR 95% CI)** | **p value** |
| --- | --- | --- | --- | --- |
| Age, years | 1 (1-19) | 0.11 | - | - |
| Male | 1 (0.55-1.8) | 0.97 | - | - |
| BMI | 1.1 (1-1.1) | **0.0003** | - | - |
| Obesity (BMI >30 kg/m<sup>2</sup>) | 2.4 (1.4-4.1) | **0.001** | 2.71 (1.33-5.51) | **0.005** |
| Hypertension | 1.6 (0.93-2.7) | 0.087 | - | - |
| Diabetes | 1.6 (0.91-2.7) | 0.11 | - | - |
| Comorbidities | 1.1 (0.93-1.4) | 0.2 | - | - |
| PaO<sub>2</sub>/FIO<sub>2</sub>, mmHg | 1 (1-1) | 0.39 | - | - |
| IMV | 0.4 (0.2-0.79) | **0.008** | 2.64 (0.99-7.06) | 0.051 |
| Prone Ventilation | 0.58 (0.32-1) | 0.7 | - | - |
| Vasoactive drugs | 0.46 (0.27-0.79) | **0.004** | 1.03 (0.47-2.36) | 0.93 |
| Creatinine, mg/dL | 1.7 (1.4-2.0) | **<0.001** | 1.44 (1.02-2.02) | **0.035** |
| Platelet/Lymphocyte ratio | 1 (1-1) | 0.39 | - | - |
| Lymphocytes 10x<sup>3</sup> mm<sup>3</sup> | 0.39 (0.19-0.78) | **0.008** | 0.78 (0.29-2.096) | 0.63 |
| Procalcitonin, ng/ml | 1 (0.99-1) | 0.3 | - | - |
| D-dimer, µg/ml | 1.1 (1.1-1.2) | **<0.001** | 1.14 (1.06-1.23) | **<0.001** |
| Troponin, pg/ml | 1(1-1) | 0.27 | -\| | - |
| C-reactive protein, mg/dL | 1 (1-1.1) | 0.054 | - | - |
| Albumin g/dL | 0.48 (0.28-0.88) | **0.007** | 0.95 (0.42-2.17) | 0.92 |

HR, Hazard ratio; CI, Confidence interval; BMI, Body mass Index; IMV, Invasive mechanical ventilation; PaO<sub>2</sub>/FiO<sub>2</sub>, Pressure arterial oxygen/Fraction inspired oxygen ratio; AKI, Acute kidney injury; Vasoactive drugs (norepinephrine and vasopressin on admission).
Admission laboratory test results were used.

## DISCUSSION

Multiple organ involvement including the liver, gastrointestinal tract and kidney have been reported in patients with COVID-19 [8]. Since information about causes leading to severe kidney disease in these patients is still limited, here we determined the factors associated with the development of AKI and explored the relation between AKI and mortality in Mexican population with severe COVID-19. We found that the group developing AKI had higher BMI values and frequency of obesity; a higher requirement of invasive mechanical ventilation and vasoactive drugs; greater levels of sCr and D-dimer on admission; and lower lymphocyte counts than the non-AKI group. The risk factors for AKI in our cohort were obesity, elevated creatinine levels and higher D-dimer levels on admission. Moreover, in-hospital mortality was significantly higher in the group with AKI, and it was particularly elevated in patients with AKI stage 3. The incidence of AKI in our cohort was 58.6%, which is higher than incidences between 5 and 7% found in China in patients with COVID-19 [8, 13, 14]. This might be partially explained by the fact that our institution is a national referral center for respiratory diseases, where mostly patients with COVID-19 severe disease are being admitted. Our study population was similar to that studied in a New York City medical center, where up to 78% of the patients developed AKI and most of them required IMV [15]

Obesity was a risk factor for AKI in our cohort and it has been reported as a common comorbidity in hospitalized patients with COVID-19 [16, 17], and as a risk factor for hospital admission and need for critical care [18]. This is particularly relevant for countries with high obesity rates, such as Mexico. In the adult Mexican population, the combined prevalence of overweight and obesity is approximately 71% [19]. A possible mechanism related to COVID-19 severity in obese persons is speculated to occur through a functional restrictive capacity of the obese lung. Moreover, chronic inflammation in obesity is apparent with an increased level of interleukin-6, adipokines and pro-inflammatory cytokines (e.g., TNF-alpha, interferon), inducing a chronic low-grade inflammatory state and impairing immune response [20, 21].

We found a higher requirement of invasive mechanical ventilation and a higher use of vasoactive drugs in patients who developed AKI, both previously reported as risk factors for AKI in patients with COVID-19 [9]. AKI, particularly when severe, is a condition that occurs among COVID-19 patients with respiratory failure [9].

We found that elevated serum creatinine on admission was a risk factor for AKI, which was previously reported in a cohort in China [8]. This means that patients with kidney involvement on admission were more likely to develop AKI. Lymphopenia was also associated with AKI in our cohort, and low lymphocyte counts have been associated with severe COVID-19 and longer hospital stay [22]. COVID-19-associated lymphopenia might derive from retention of lymphocytes in the lung. Also, lymphocytes express the angiotensin-converting enzyme 2 (ACE2) receptor on their surface [23]. Thus, SARS-CoV-2 infection may directly induce lysis of these cells. In addition, elevated levels of pro-inflammatory cytokines, may promote lymphocyte apoptosis.

One additional risk factor for AKI found here was D-dimer elevation. This molecule is a product of cross-linked fibrin degradation and is a sensitive marker of thrombosis and coagulation activation [24]. Elevated D-dimer level has been consistently reported in patients with COVID-19 [25, 26], and its gradual increase during disease course is particularly associated with disease worsening [27].

The main limitation of our study is its retrospective design. Also, the number of patients included in the study is limited. Another limitation is that we retrieved information during hospitalization, but a longer observation period would have provided additional information regarding the clinical outcome and the impact of AKI in the population studied. That is, we were not able to report the proportion of individuals developing chronic renal disease in the group with AKI. The lack of pre-hospital baseline sCr measurements was also a study limitation because baseline sCr values were an estimation. Since we could not assess the baseline renal status, we could not explore whether chronic kidney disease itself is a risk factor for AKI in the context of COVID-19. Finally, our study was conducted in a national referral center for respiratory diseases receiving disproportionately more patients with poor outcomes, and this represents a potential source of referral bias.

### Conclusion

AKI was common in our cohort of patients with severe COVID-19 and it was associated with mortality. The risk factors for AKI were obesity, elevated creatinine levels and higher D-dimer levels on admission.

## DISCLOSURE

All the authors declared no competing interests.

## Data Availability

We have the approval of the approved ethics committee and the database of our protocol.

